# Current and lagged associations of meteorological variables and *Aedes* mosquito indices with dengue incidence in the Philippines

**DOI:** 10.1101/2023.08.21.23294355

**Authors:** Estrella I. Cruz, Ferdinand V. Salazar, Ariza Aguila, Jennifer Ramos, Richard E. Paul

**Author notes:** These authors contributed equally to this work (Estrella I. Cruz, Ferdinand V. Salazar).

## Abstract

**Background:** Dengue is an increasing health burden that has spread throughout the tropics and sub-tropics. There is currently no effective vaccine and control is only possible through integrated vector management. Early warning systems to alert potential dengue outbreaks are currently being explored but despite showing promise are yet to come to fruition. This study addresses the use of meteorological variables for predicting both entomological indices and dengue incidences and assesses the added value of additionally using entomological indices for predicting dengue incidences.

**Methodology/Principal Findings:** Entomological surveys were carried out monthly for 14 months in six sites spread across three environmentally different cities of the Philippines. Meteorological and dengue data were acquired. Non-linear generalized additive models were fitted to test associations with the meteorological variables and both entomological indices and dengue cases. Rain and the diurnal temperature range (DTR) contributed most to explaining the variation in both entomological indices and number of dengue cases. DTR and minimum temperature also explained variation in dengue cases occurring one and two months later. The number of adult mosquitoes did associate with the number of dengue cases, but contributed no additional value for predicting dengue cases.

**Conclusions/Significance:** The use of meteorological variables to predict future risk of dengue holds promise. The lack of added value of using entomological indices confirms several previous studies and given the onerous nature of obtaining such information, more effort should be placed on improving meteorological information at a finer scale to evaluate efficacy in early warning of dengue outbreaks.

**Author summary:** Dengue is a widespread mosquito-borne disease. Mosquitoes are sensitive to temperature and rainfall and hence there have been efforts to identify such variables for predicting dengue outbreaks. Several mosquito indices are measured routinely by national surveillance systems, but which vary considerably in their success of predicting dengue outbreaks. This study explored the current and lagged associations of meteorological variables with mosquito indices and dengue incidence. Associations of mosquito indices with dengue were also explored. Rain and the diurnal temperature range (DTR) contributed most to explaining the variation in both entomological indices and number of dengue cases. DTR and minimum temperature also explained variation in dengue cases occurring one and two months later. Mosquito indices did not provide any additional predictive power of dengue incidences. Given the onerous nature of measuring mosquito indices, advanced warning systems might be improved using meteorological variables measured at finer scales than that traditionally available.

## Introduction

Dengue is a rapidly spreading mosquito-borne infectious disease caused by any of the four serotypes of the dengue virus (DENV 1-4). Despite the known underestimation of its real global burden [1], it is estimated that dengue incidence has increased 30-fold over the last few decades [2]. The disease is endemic in over 100 countries and more than 3.5 billion people are at risk of DENV infection [3,4]. Southeast Asia and the Western Pacific have historically been and are still among the most affected places [5-8].The public health significance of dengue in the Philippines has continued since its initial discovery in 1954 to date [9]. The exceptionally high numbers of reported infections incidence and dengue-associated mortality cases in a span of five years, from 2010 to 2014, claiming at least 812,525 victims and resulting in nearly 3,500 deaths, illustrates the high burden in the population.

Several environmental and socio-economic factors such as weather, urbanisation and globalisation have been associated with the spread of dengue [10-14] and this spread depends on the presence and abundance of the arthropod vector [15]. Moreover, in light of recent anthropogenic environmental impact, there has been a growing concern over global climate change as a potential factor increasing the risk of dengue through both the increased distribution of the mosquito vector species and the increased capacity for the vector to transmit the virus (the vectorial capacity) [16, 17].

DENV transmission is shaped by climatic conditions such as temperature and rainfall [18]. *Aedes spp*. reproductive and feeding behaviours, as well as viability of the species, depend, at least partly, on these environmental variables and thus have been widely studied. Mosquito abundance is partially regulated by rainfall by providing breeding sites and triggering egg hatching [19]. Temperature influences the life span of the mosquito vector and has a direct effect on developmental and feeding rates [19-21]. In addition to this, DENV replication inside the vector also increases in warmer temperatures [21, 22].

Having no fully effective vaccine to prevent infection, or drugs to treat the infection, vector control is still the main strategy to prevent transmission of DENV. Vector monitoring and surveillance is an evidence-based, analytic approach to better understand the mosquito vector population dynamics and virus transmission. Combined with a community-based strategy, which requires direct and immediate action in the community using Integrated Vector Management practices and Communication for Behavioral Impact approaches, vector surveillance offers the best defense against the vector and disease [23-25]. In urban settings, the major vector species is *Aedes aegypti*, which has adapted to the peridomestic environment, ovipositing in man-made artificial containers. Several *Aedes* indices, such as the House, Container, Breteau and Pupal indices, have been proposed for monitoring strategies to predict risk of dengue transmission. However, to date there is little consensus on the appropriate threshold values to trigger a mosquito control response and their general utility to avert an outbreak [26-30].

Identifying the factors that are implicated in DENV transmission and being able to forecast the onset of dengue outbreaks in endemic areas would provide the opportunity to be prepared and implement timely responses to decrease the burden of dengue in human populations. Efforts have been made to predict the incidence of infection using meteorological data [31, 32]. However, our understanding remains poor over the relative contributions of meteorological variables to these indices in the context of urban settings where the microenvironment and the abundance of artificial oviposition sites impact mosquito abundance [17]. While other prevention and control measures for dengue are being developed, understanding the relationship between the risk of disease and meteorological factors in an urban setting for elaborating early warning systems remains of key interest.

The aim of this study was to expand the current knowledge on the associations between different meteorological variables and both the *Aedes* indices and the incidence of dengue and to identify the meteorological and mosquito indices that provide advanced warning of dengue outbreaks in three representative cities in the Philippines.

## Methods

### Study sites (Figure 1)

The study was carried out in six barangays, of which two were in each of the three study cities, Manila and Muntinlupa on the major island of Luzon and Puerto Princesa on Palawan island. In the highly densely populated area of Manila, two barangays were selected: Sampaloc, Manila (14° 36’ 39.708“N 120° 59’ 46.4496“E) covering 7.90 km^2^ with a population of 388,305 and Tambunting, Santa Cruz District, Manila (14°37’46.3“N 120°59’01.5“E) covering 3.07 km^2^ with a population of 126,735. In the more recently urbanized and less dense city of Muntinlupa, two barangays were selected: Cupang (<u>14°25′53.4″N 121°2′55″E</u>) covering 5.37 km^2^ with a population of 57,013 inhabitants and Putatan (<u>14°23′54.12″N 121°2′10.96″E</u>) covering 6.75 km^2^ with a population of 99,725. On the island of Palawan, which has a much lower degree of urbanization and population density, two urban barangays were chosen within Puerto Princesa, a coastal city covering 2,381 km^2^ with a population of 307,079. Two selected barangays were: San Miguel (09°44’39.48”N 118°44’44.16”E), considered an urban barangay with a population of 21,157 and San Pedro (09°45’19.44”N 118°45’2.52”E) a neighboring urban barangay with a population of 25,909.

**Figure 1.**
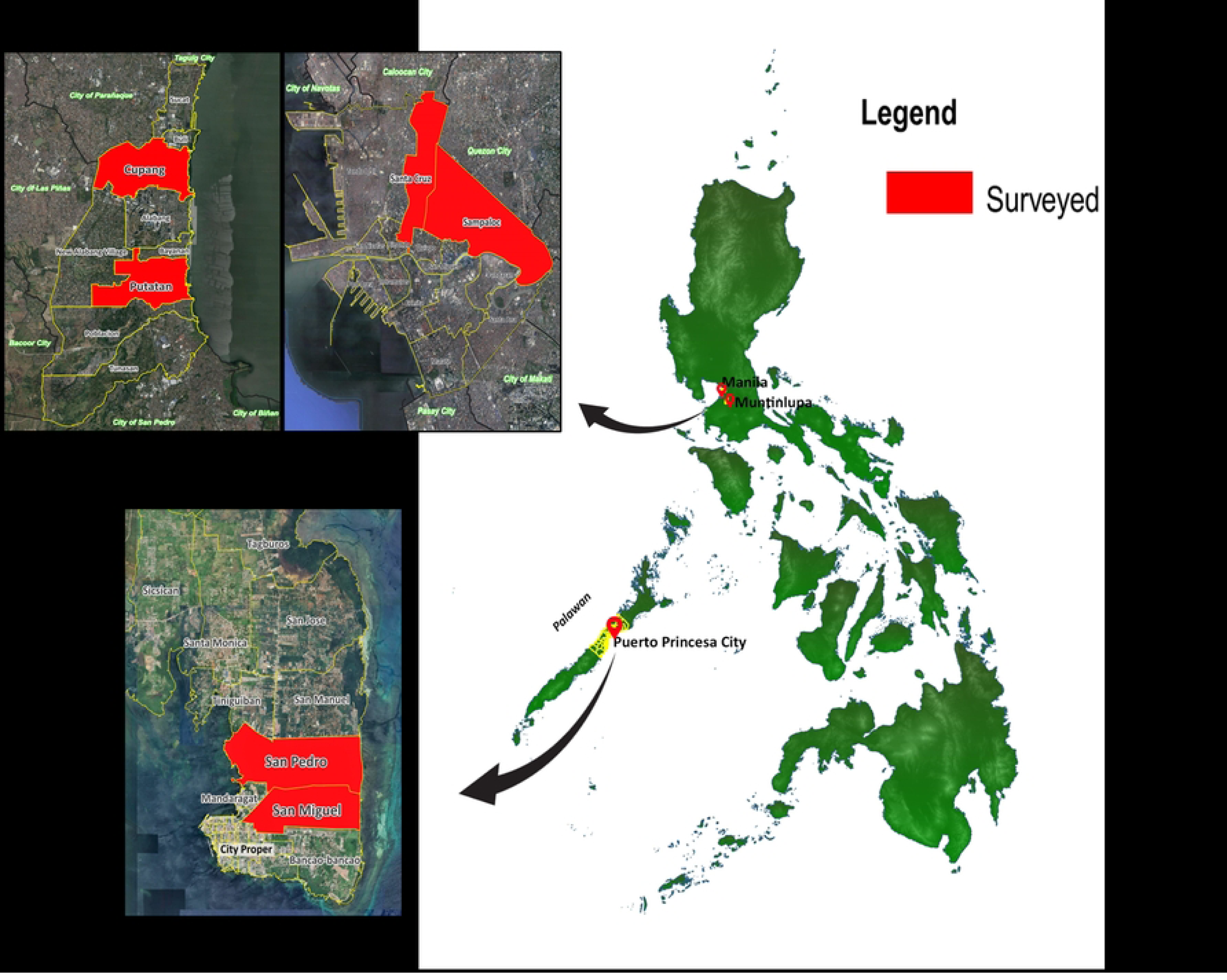
Study Sites.

### Entomological, meteorological and dengue case data

Entomological surveys were carried out monthly from November 2014 until December 2015, thus 14 months duration. Dengue case data were available monthly from October 2014 until December 2015. Meteorological data were available on a daily basis, provided by the Philippine Atmospheric, Geophysical and Astronomical Services Administration (PAG-ASA) from meteorological stations closest to the areas under study. Meteorological data included rain, relative humidity and temperature. From these data the cumulative monthly rain (mm), the monthly mean daily rain (mm), the monthly mean daily Relative Humidity (%), the monthly mean daily minimum, maximum and mean temperatures (°C) and the monthly mean daily Diurnal Temperature Range (DTR, °C) were calculated.

Mosquito indices: Immature mosquito life stages surveillance was conducted in 100 households per selected barangay wherein all water-holding containers were inspected for presence of immature stages of mosquito. Larvae and pupae found in the containers were all collected by pipetting. Specimens from each positive container were transferred in separate ice plastic bags for transport to the laboratory for rearing and identification. The number of water-holding containers (artificial or natural) with or without cover that were found indoors or outdoors, together with the number of containers found positive for larvae and/or pupae were recorded. The number of people who slept inside the house the previous night was also noted. The following indices were used: the Container Index (CI, number of water-filled containers positive for larvae or pupae), the House Index (HI, number of houses found positive for larvae or pupae), the Breteau index (BI, the number of containers positive divided by the number of houses visited), the Pupal Index (PI, the number of pupae divided by the number of houses visited x 100), the number of pupa per person (PPI, the number of pupae divided by the total population of the inspected households). In addition to these immature *Aedes* spp. mosquito stage indices, adult mosquitoes were captured in the houses using a sweep net each month. Adult mosquitoes were collected by circumnavigating the internal periphery of the house from the front door to the different rooms while continually moving the net in a figure of eight at 90° or at 180° targeting known resting places of adult mosquitoes. These areas include areas under the beds, hanging clothes, under sink, comfort rooms, closet, dark cool room of the house shoe racks, and outdoors such as vegetation, bushes, tree hook and plants. Trapped mosquitoes were transferred to Styrofoam cups with the use of sucking tube. All collected mosquitoes were identified morphologically to species in the laboratory. In addition to the monthly adult *Aedes* spp. mosquito count, a cumulative two-monthly count was calculated.

### Statistical analyses

To test the association of the meteorological variables with the entomological indices, a Generalized Additive model (GAM) was fitted to account for any non-linearity in the relationships. For indices concerning proportions (i.e. House Index and Container Index), a logistic regression was fitted. For the indices concerning counts, a loglinear regression was fitted. First all variables were fitted in a univariable analysis and those achieving a P-value less than 0.25 were fitted in the multivariable analysis. Barangay was nested within City. The multivariable analysis proceeded by step-wise elimination of non-significant variables until a final adequate model containing only significant variables was achieved. Because many of the variables were strongly correlated (i.e. the min, mean and max temperature variables), and thus led to collinearity and potentially spurious non-significance, models were refitted with removal and replacement to identify which of the variables were the most strongly associated (based on % variance explained). For the analyses of the association of the meteorological variables with the number of dengue cases, a similar approach was performed but including the natural log of the population of each barangay as an offset. When using the entomological indices as explanatory variables, the data were transformed either by using an arcsine transformation for the proportion/percentage indices or by standardization for the count indices (subtraction of the mean and division by the standard deviation). A dispersion parameter was estimated to account for any over-dispersion of the data. All analyses were conducted in Genstat version 22 [33].

## Results

### Description of city specific meteorological variables, dengue cases and mosquito catches

Rainfall showed distinct seasonality, with the dry season from January to April and the peak rainy season from July to October (Fig. S1). There was little variation among the three cities, with the monthly mean of the daily rainfall varying from 0.014-14.35 mm in Muntinlupa, 0-9.24 mm in Manila and 0-10.16 mm in Palawan. Relative humidity was consistently lower from February to June in all three cities (Fig. S2). Palawan varied the least over the year (73.7-82.5%) as compared to Manila (61.8-84.6%) or Muntinlupa (65.8-81.0%). Maximum temperatures reached a peak from April to June (Maximum 35°C in Manila, 34-35°C in Muntinlupa and 32-33°C in Palawan) (Fig. S3). Year round temperatures varied little with minimum temperatures oscillating around 24-27°C and mean temperatures around 25-31°C (Fig. S4 and Fig. S5). Temperatures varied little across the three cities but were generally less variable in Palawan as compared to Manila and Muntinlupa. The same was seen for the Diurnal Temperature Range with a peak from March to June (8-9°C) and higher values in Manila and Muntinlupa than Palawan (6-9°C vs. 6-8°C) (Fig. S6).

Over the study period, dengue cases were concentrated from July-December 2015. Overall, there were 343 and 369 in Tambunting and Sampaloc (Manila), respectively, 76 and 240 in Cupang and Putatan (Muntinlupa), respectively and 188 and 110 in San Miguel and San Pedro (Palawan). Dengue Incidence rates (IR) per 10,000 individuals were initially low (0-5.2 / 10000) until July when an epidemic occurred, notably in San Miguel, Palawan, with IRs ranging from 9.0-28.4 / 10000 (Figure 2).

**Figure 2.**
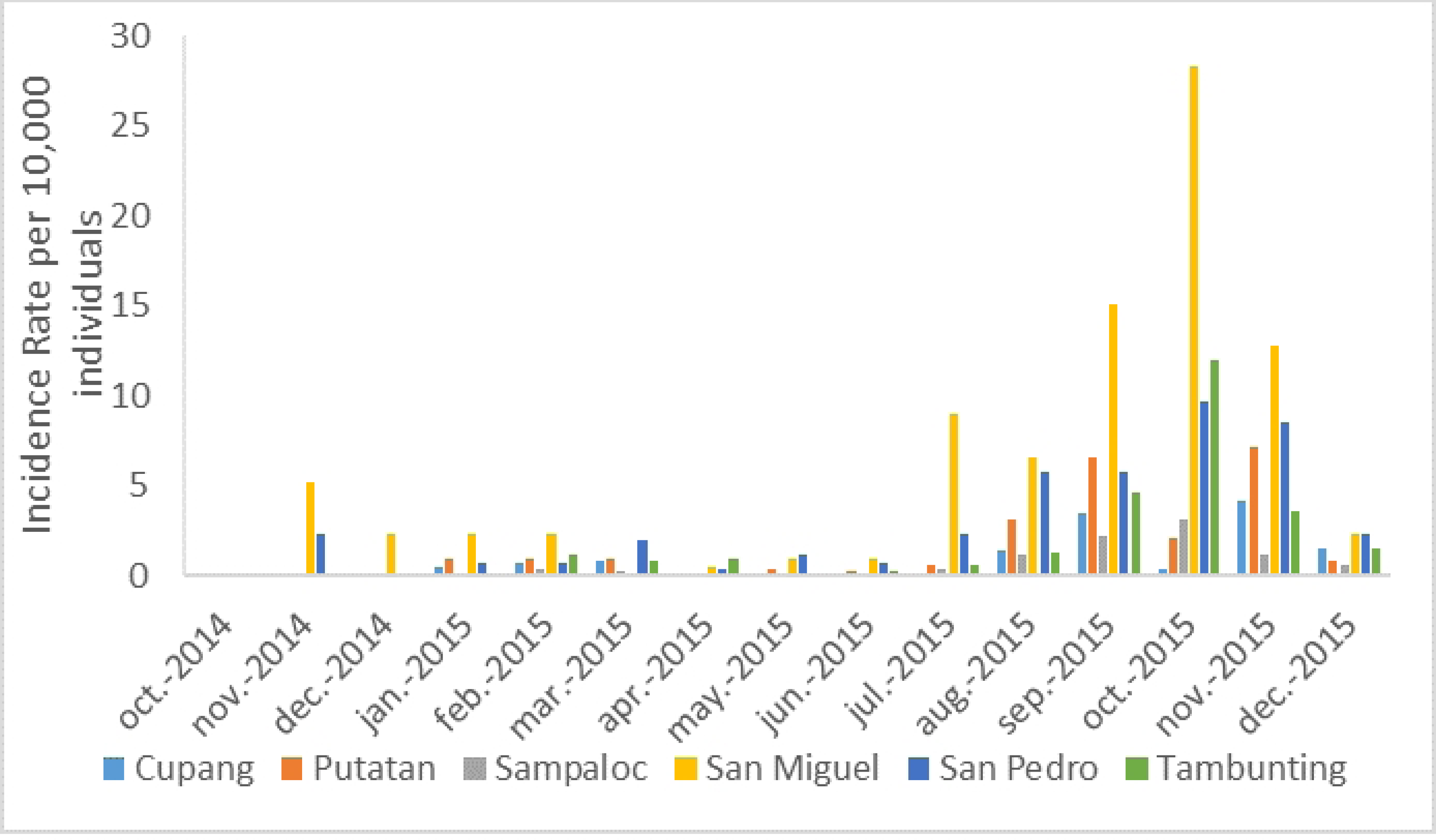
Dengue Incidence Rate per 10,000 individuals in the six study sites.

Overall 85.4% of the immature mosquito stages were *Aedes aegypti*, 12.3% *Aedes albopictus* and 2.4% *Culex* or other genus spp. In the Luzon study sites (Manila and Muntinlupa), 96.8% were *Ae. aegypti*, 1.9% were *Ae. albopictus* and 1.4% *Culex* or other genus spp. In the two Palawan study sites, 79.5% were *Ae. aegypti*, 17.5% *Ae. albopictus* and 2.9% *Culex* or other genus spp. For the adult mosquito catches, 50.5% were *Ae. aegypti*, 0.2% *Ae. albopictus* and 49.3% *Culex* or other genus spp. For the subsequent calculation of mosquito indices, only *Aedes* spp. numbers were used. Because of the differing relative abundance of *Ae. albopictus* in Palawan, for the association of mosquito indices with weather variables we first analyzed all the sites together and then the two Palawan and four Luzon sites alone.

### Association of meteorological variables with mosquito indices

The House indices (HI) were very high, reaching 50% in some sites in the months following the onset of the rains in July (Mean Rain Figure S1 & House Index Fig. S7). Indices were highest in San Pedro and San Miguel in Palawan and Putatan in Muntinlupa, the latter being exceptionally high as compared to the other sites during the dry season, reaching 20%. Association analyses revealed that in addition to the variation explained by location (27.2% variance explained), meteorological variables explained 57.4% of the observed variation in HI (Table 1). Cumulative monthly rainfall and mean daily rainfall showed the same relationship with HI, where HI increased gradually before reaching a plateau at ∼150mm for Cumulative monthly rain and ∼5mm for mean daily rain per month (See Supplementary Figure S8 for Fitted plot of HI against Cumulative Rain). The monthly mean of the Diurnal Temperature range exhibited an increase in HI to a peak at 6-6.5°C before a subsequent decline (Figure 3).

**Figure 3.**
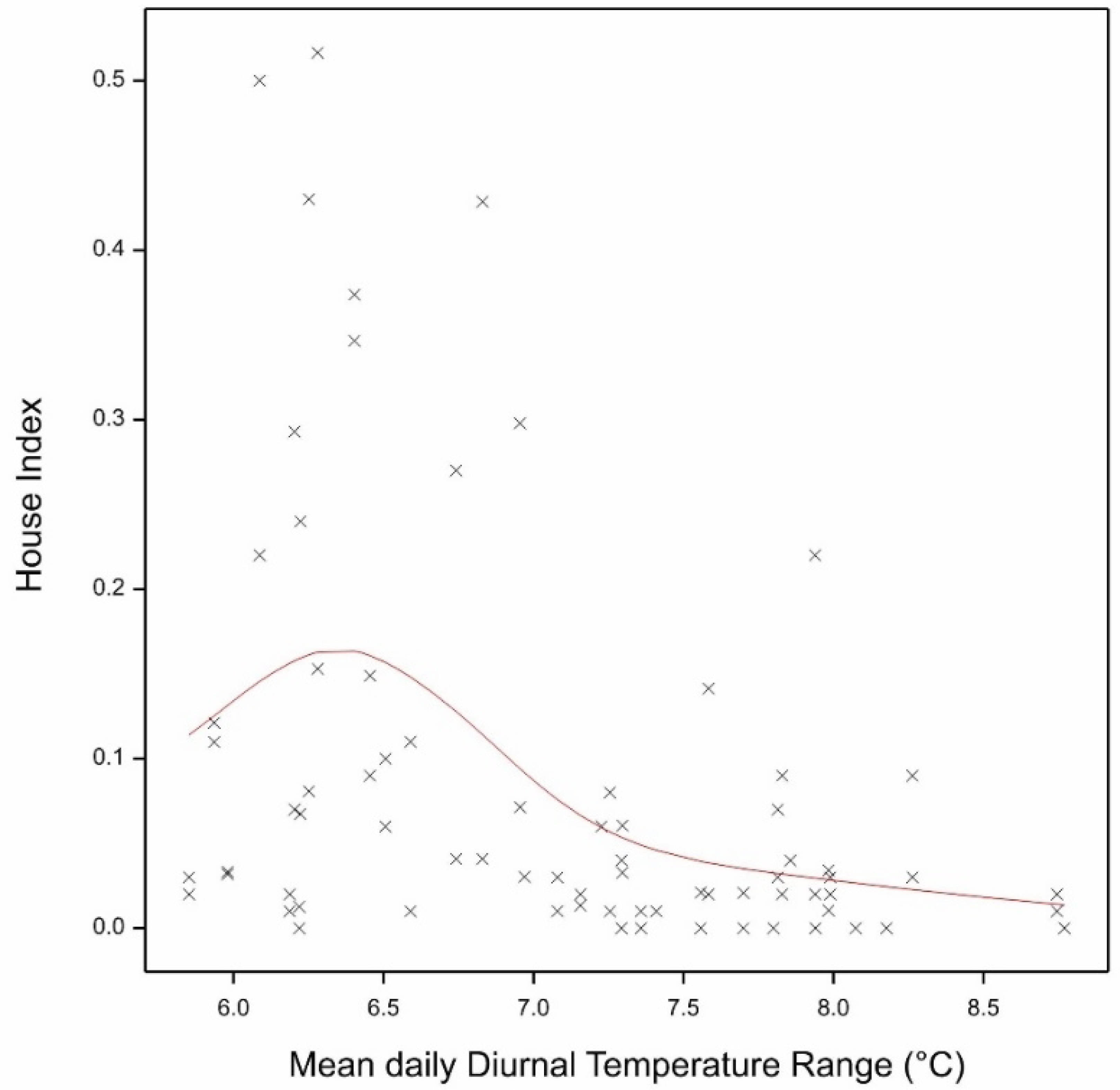
The fitted logistic regression of the House Index (here shown as a proportion) against the Diurnal Temperature range. The red line shows the fitted model in the GAM.

**Table 1.** Association of meteorological variables with Mosquito indices.

|  |  | P value | variance explained (%) |
| --- | --- | --- | --- |
| House Index | Cumulative Rain (mm) | <.001 | 13.37 |
|  | Mean DTR (°C) | <.001 | 20.71 |
|  | Mean Rain (mm) | <.001 | 15.10 |
|  | Mean RH (%) | <.001 | 5.01 |
|  | Min. Temp. (°C) | 0.01 | 3.22 |
| Container Index | Cumulative Rain (mm) | <.001 | 15.25 |
|  | Mean DTR (°C) | <.001 | 20.19 |
|  | Mean Rain (mm) | <.001 | 6.45 |
|  | Mean RH (%) | 0.001 | 5.39 |
|  | Min. Temp. (°C) | *0.016* | 3.34 |
| Breteau Index | Cumulative Rain (mm) | <.001 | 14.94 |
|  | Mean DTR (°C) | <.001 | 25.16 |
|  | Mean Rain (mm) | <.001 | 14.57 |
|  | Mean RH (%) | 0.001 | 4.14 |
|  | Min. Temp. (°C) | 0.002 | 3.82 |
| Pupal Index | Cumulative Rain (mm) | *0.009* | 6.90 |
|  | Mean DTR (°C) | <.001 | 17.38 |
|  | Mean Rain (mm) | *0.009* | 6.85 |
|  | Mean RH (%) | *0.007* | 7.30 |
|  | Min. Temp. (°C) | *0.011* | 6.61 |
|  | Max. Temp. (°C) | *0.043* | 3.58 |
| Pupa per person | Cumulative Rain (mm) | *0.007* | 6.94 |
|  | Mean DTR (°C) | <.001 | 18.03 |
|  | Mean Rain (mm) | *0.013* | 6.14 |
|  | Mean RH (%) | 0.006 | 7.19 |
|  | Min. Temp. (°C) | *0.01* | 6.44 |
|  | Max. Temp. (°C) | *0.033* | 3.78 |
| Adults | Cumulative Rain (mm) | <.001 | 36.26 |
|  | Mean DTR (°C) | 0.006 | 4.76 |
|  | Mean Rain (mm) | 0.006 | 4.82 |
|  | Mean RH (%) | <.001 | 7.22 |
|  | Min. Temp. (°C) | <.001 | 14.42 |
|  | Max. Temp. (°C) | 0.003 | 4.52 |

P values in italics are those above the Bonferroni corrected P threshold for multiple tests (7 meteorological variables, plus one multivariable analyses, thus 8 models per index; P=0.0063).

Container indices (CI) were substantially lower, generally less than 5%, except reaching a maximum of 17% in San Pedro following the rains (Figure S9). Once again, CIs were higher following the rains in San Pedro, Palawan and during the dry season in Putatan, Muntinlupa. The same relationships were observed with the meteorological variables as for HIs, explaining 50.6% of the variation in CIs (Table 1).

Breteau indices in the two sites in Manila never exceeded a BI>=5 (Figure S10). In Cupang, Muntinlupa, BI> 5 occurred on three occasions but Putatan was consistently higher than 5 and again showed aberrantly high values (10-23) during the dry months. Values in the two sites in Palawan were consistently very high throughout the year except for the dry season months (February to June); values reached over 100 in San Pedro and 9-23 in San Miguel during the dengue epidemic period from July onwards in 2015. The same associations and relationships with meteorological variables were observed, explaining 63% of the variation in BI (Table 1).

The Pupal and Pupa per person indices (PI and PPI) were highly variable ranging from 0 to over 100% (Figure S11 and Figure S12). With the exception of Putatan, which showed the aberrantly high pupal indices in the dry season months, PIs were generally zero during the dry season and then rapidly increased in the months following the rains. The same meteorological associations were observed, albeit generally weaker than those observed for the HI, CI and BI. However, these variables still explained 49% of the observed variation in PI and PPI (Table 1).

In contrast to these immature stage indices, adult *Aedes spp.* were less abundant in the two Palawan sites and most abundant in Putatan, especially during the dry season (Figure S13). The relationship of the meteorological variables with the number of adult *Aedes* spp. mosquitoes also differed (Table 1). There was a strong association with Cumulative rain, with progressively increasing numbers of *Aedes* spp. adults caught with increasing rain (Figure S14). There was a distinct non-linear relationship with minimum temperature, with a peak at 25-26°C (Figure S15). The strength of the relationships with the other meteorological variables was much weaker, but combined overall explained 72% of the observed variation.

Analysis of the two sites on Palawan separately from the Luzon study sites revealed a similar but simpler series of relationships with the meteorological variables. For HI, CI and Breteau, Cumulative rain and DTR were significantly associated (Cumul. Rain: % variance explained 43.8%, 39.8% and 46.9% for HI, CI and Breteau respectively; DTR explained 13.1%, 14.2% and 17.3% of the variance for HI, CI and Breteau respectively). For the Pupal Index, Cumulative rain and Relative Humidity explained 28.4% and 31.0% of the variance. For the PPI, Cumulative rain, DTR and Relative Humidity explained 26.2%, 17.5% and 20.4% of the variance. Finally, for the adults, Cumulative rain and DTR were significantly associated, explaining 44.3% and 27.2% of the variance.

### Association of meteorological variables with lagged mosquito indices

The association analyses were then repeated for lag periods of one to four weeks. Monthly values of the meteorological variables were thus calculated for the weeks preceding the mosquito index counts. With the exception of the Pupal Index, lagged associations explained less of the observed variation in the mosquito indices (Table S1). For the Pupal Index, variables lagged by two weeks explained 52.9% of the variation as compared 48.6% for the unlagged associations. In both cases there was a notable decrease in PI with increasing DTR (Figure S16) and a significant increase in PI with increasing Cumulative rain (Figure S17).

### Association of meteorological variables with dengue cases

There were several associations of meteorological variables with the number of dengue cases during the same month (Table 2). There was a gradual increase in the number of cases with increasing rain (both Cumulative and monthly daily mean) (Figure 4) and Relative Humidity. Dengue cases peaked at a maximum temperature of 32-33°C, a minimum temperature of 25-26°C and a DTR of 6.5-7°C (Figure S18 for the association with DTR). Overall the meteorological variables explained 60.4% in the variation of observed cases. Assessing the association with dengue cases the following one or two months explained less of the variation (38.9% and 39.3% respectively) and the rain variables became non-significant. However, the associations with DTR, RH and especially Minimum temperature remained significant and followed the same form (Figure 5).

**Table 2.**
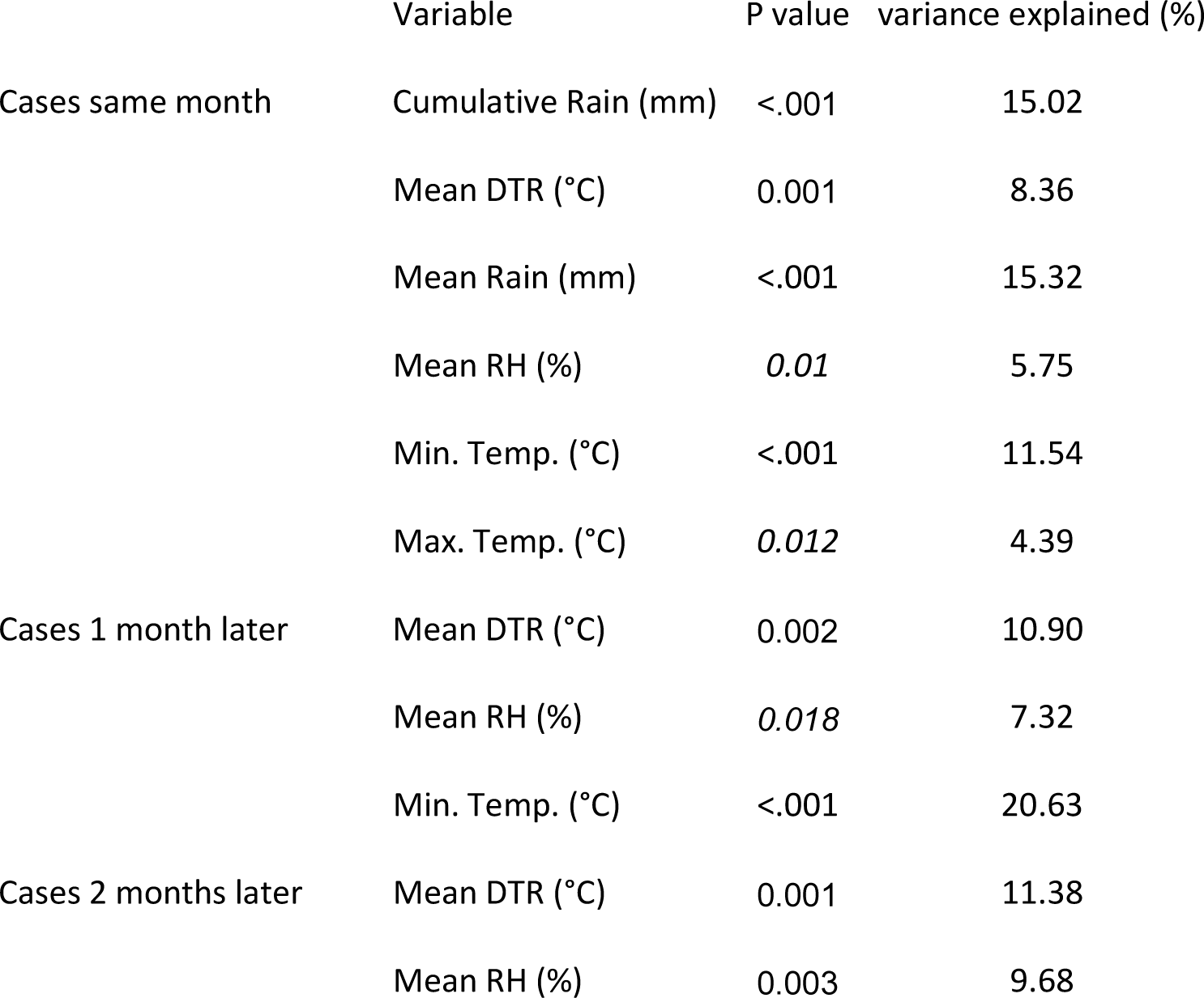
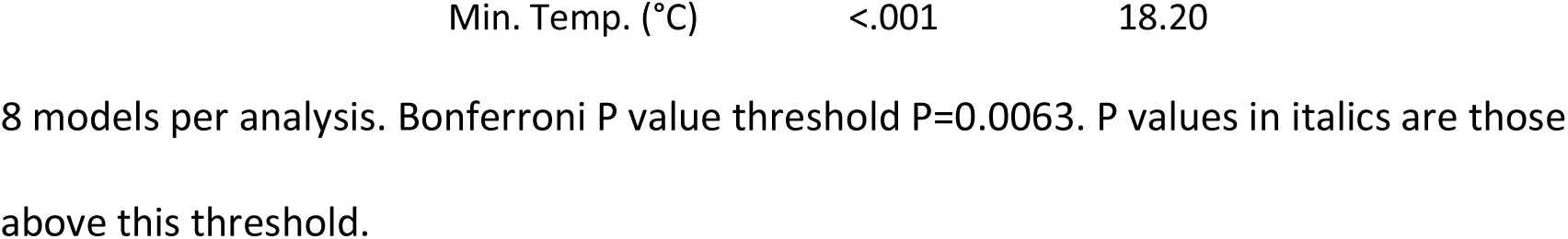
Association of meteorological variables with dengue cases.

|  | Variable | P value | variance explained (%) |
| --- | --- | --- | --- |
| Cases same month | Cumulative Rain (mm) | <.001 | 15.02 |
|  | Mean DTR (°C) | 0.001 | 8.36 |
|  | Mean Rain (mm) | <.001 | 15.32 |
|  | Mean RH (%) | *0.01* | 5.75 |
|  | Min. Temp. (°C) | <.001 | 11.54 |
|  | Max. Temp. (°C) | *0.012* | 4.39 |
| Cases 1 month later | Mean DTR (°C) | 0.002 | 10.90 |
|  | Mean RH (%) | *0.018* | 7.32 |
|  | Min. Temp. (°C) | <.001 | 20.63 |
| Cases 2 months later | Mean DTR (°C) | 0.001 | 11.38 |
|  | Mean RH (%) | 0.003 | 9.68 |

|  |  |  |
| --- | --- | --- |
| Min. Temp. (°C) | <.001 | 18.20 |

8 models per analysis. Bonferroni P value threshold $P=0.0063$ . P values in italics are those above this threshold.

**Figure 4.**
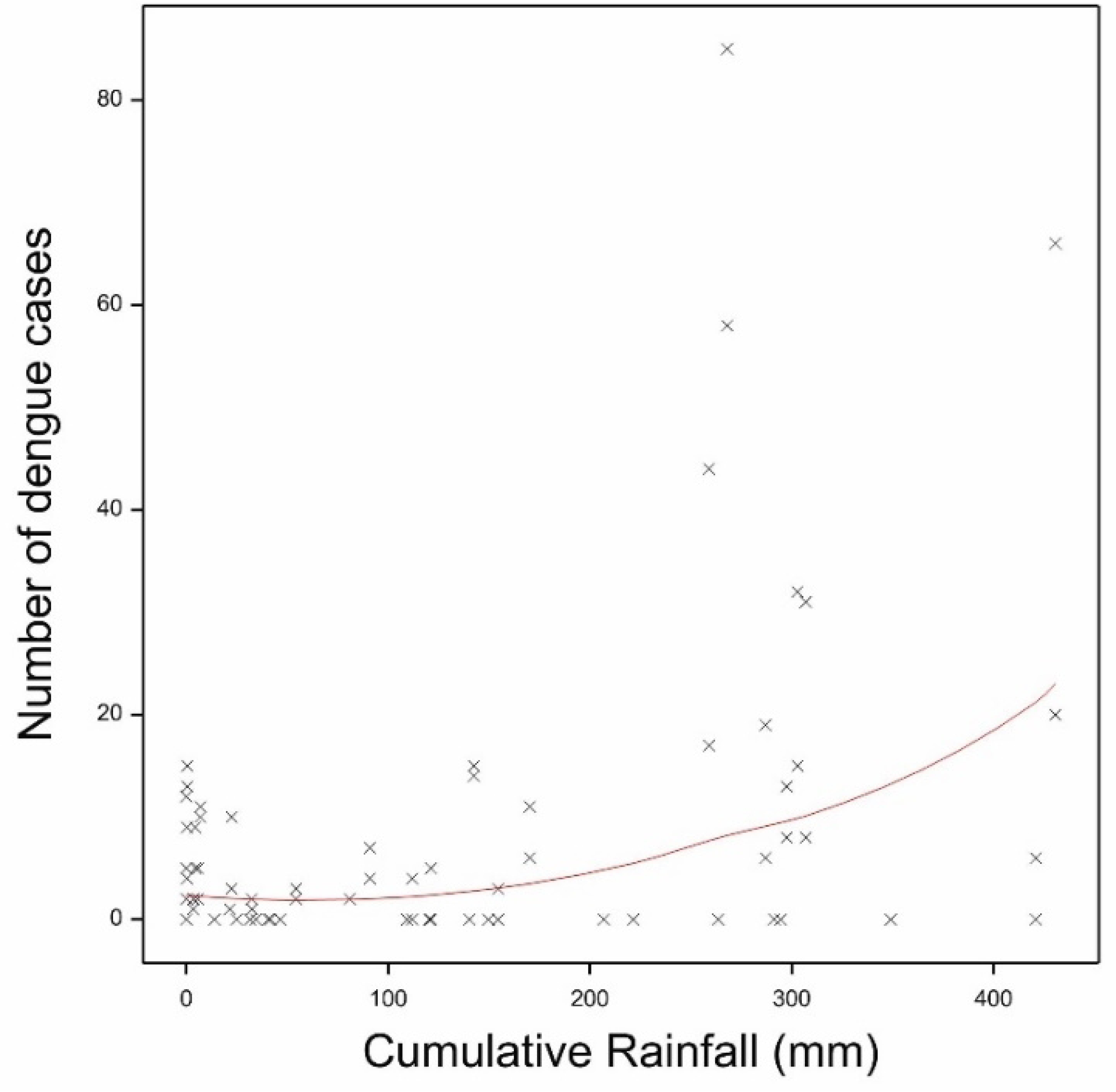
Fitted model of the association between Cumulative rain and dengue cases in the same month. Shown are the observed data (crosses) and the fitted model (red line).

**Figure 5.**
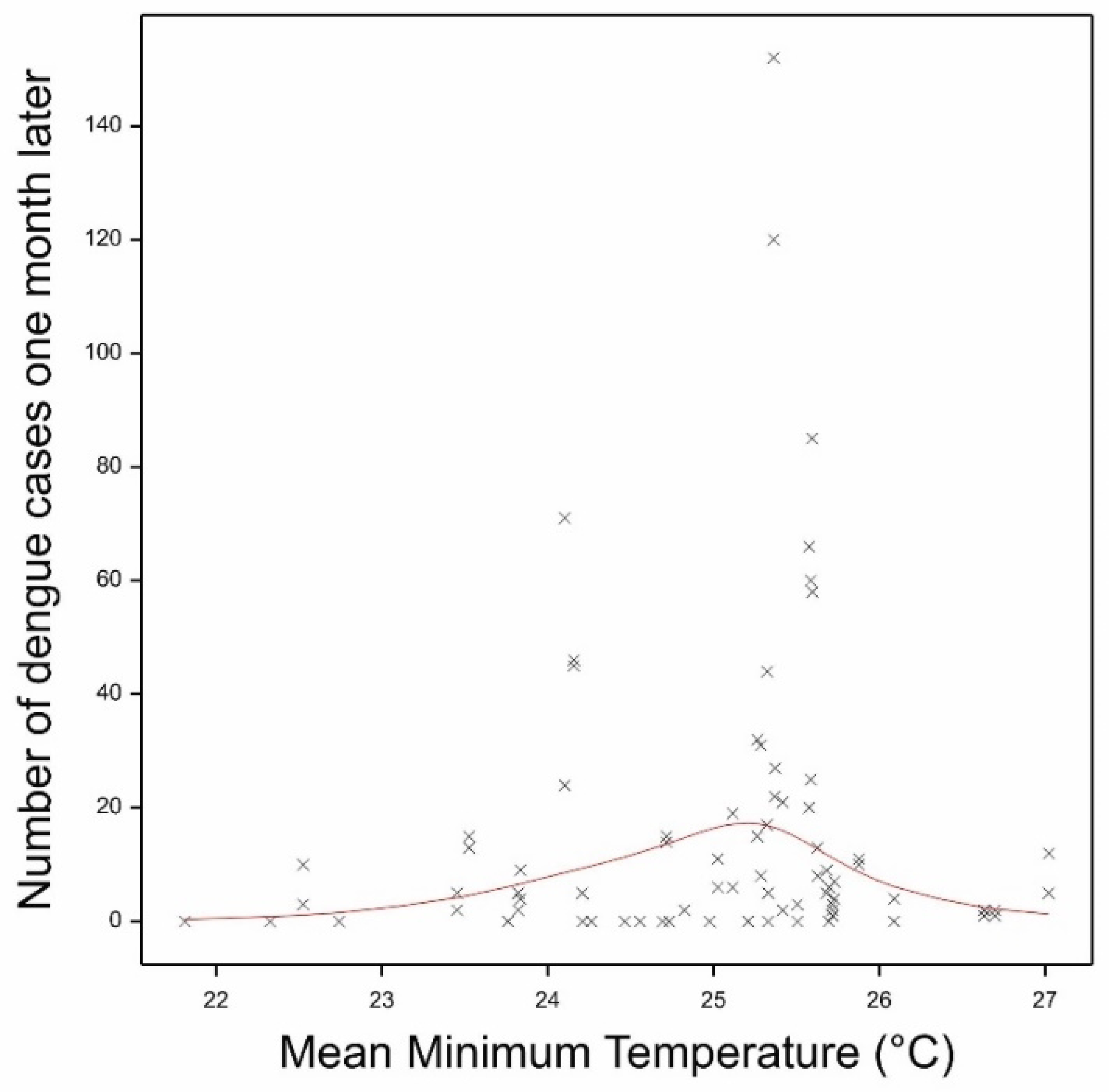
Mean Daily Minimum Temperatures and the number of dengue cases one month later.

Analyzing the two Palawan sites independently of the four Luzon sites revealed a much improved model fit, with Cumulative rain, DTR and Relative Humidity explaining 71.3% of the variation in the number of dengue cases in the same month. Moreover, Cumulative rain, DTR and Mean maximum temperatures explained 73.9% and 71.7% of the variation in cases one and two months later respectively. The model fit for the association of the meteorological variables with the number of dengue cases two months later is shown in Figure 6. 10.2% of the variation was accounted for by the site (San Miguel vs. San Pedro). Likewise, when analyzing the four sites on Luzon independently of Palawan gave a much improved model fit, explaining 69.5%, 48.1% and 77.9% of the variation in dengue cases the same month and one and two months later. Minimum temperatures and DTR contributed the most. The model fit for the association of the meteorological variables with the number of dengue cases two months later is shown in Figure 7. However, when analyzing Muntinlupa and Manila separately, there was no improved model fit from when analyzing all four Luzon sites together.

**Figure 6.**
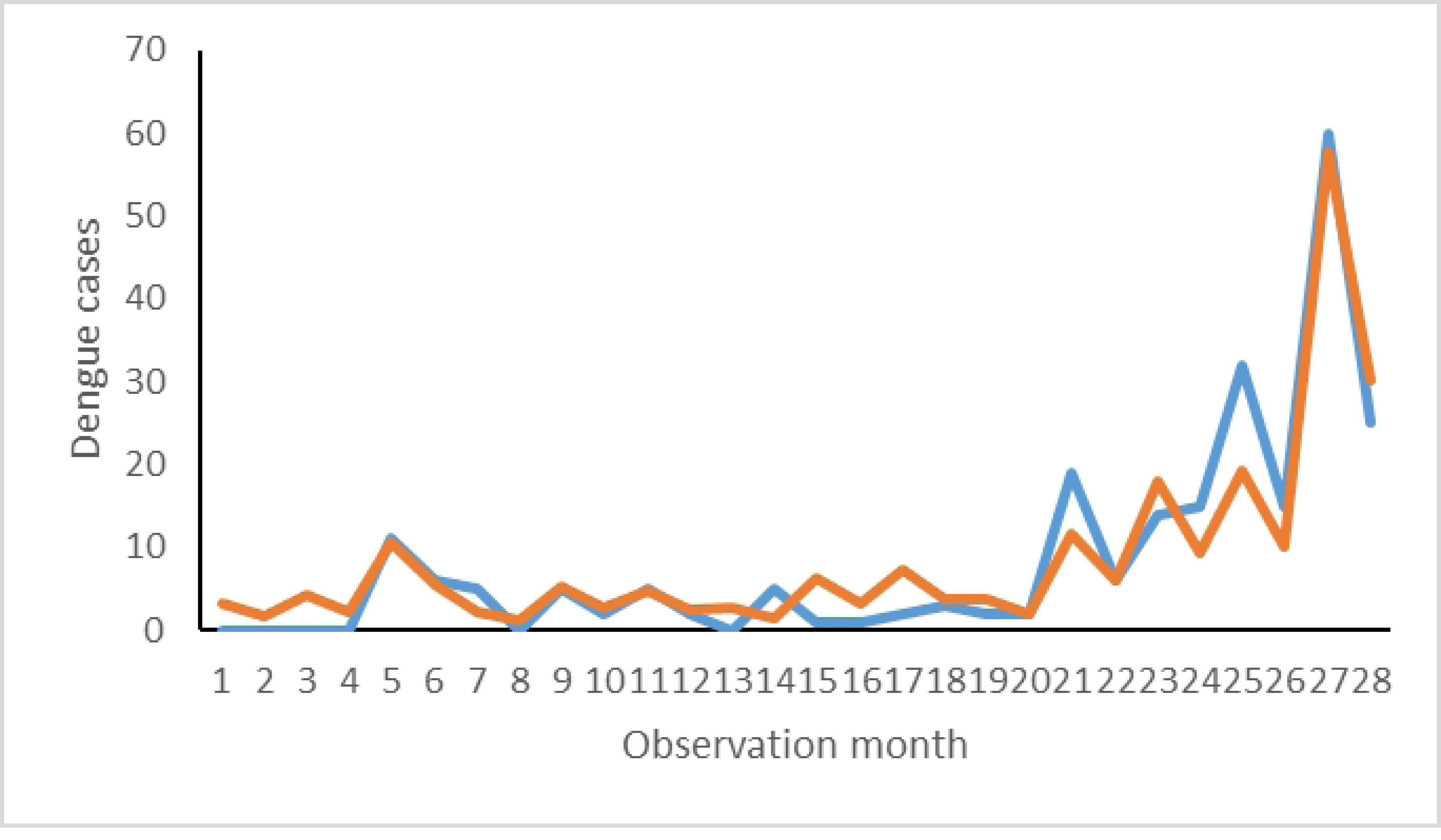
Fitted model of meteorological variables with the observed number of dengue cases two months later in the two study sites on Palawan. Observed values shown by the blue line and fitted values by the orange line. The X-axis is the observation month for each of the two study sites starting from the beginning of the study until the end. Thus months 1 & 2 will be the first observation months of San Miguel and San Pedro respectively and so forth.

**Figure 7.**
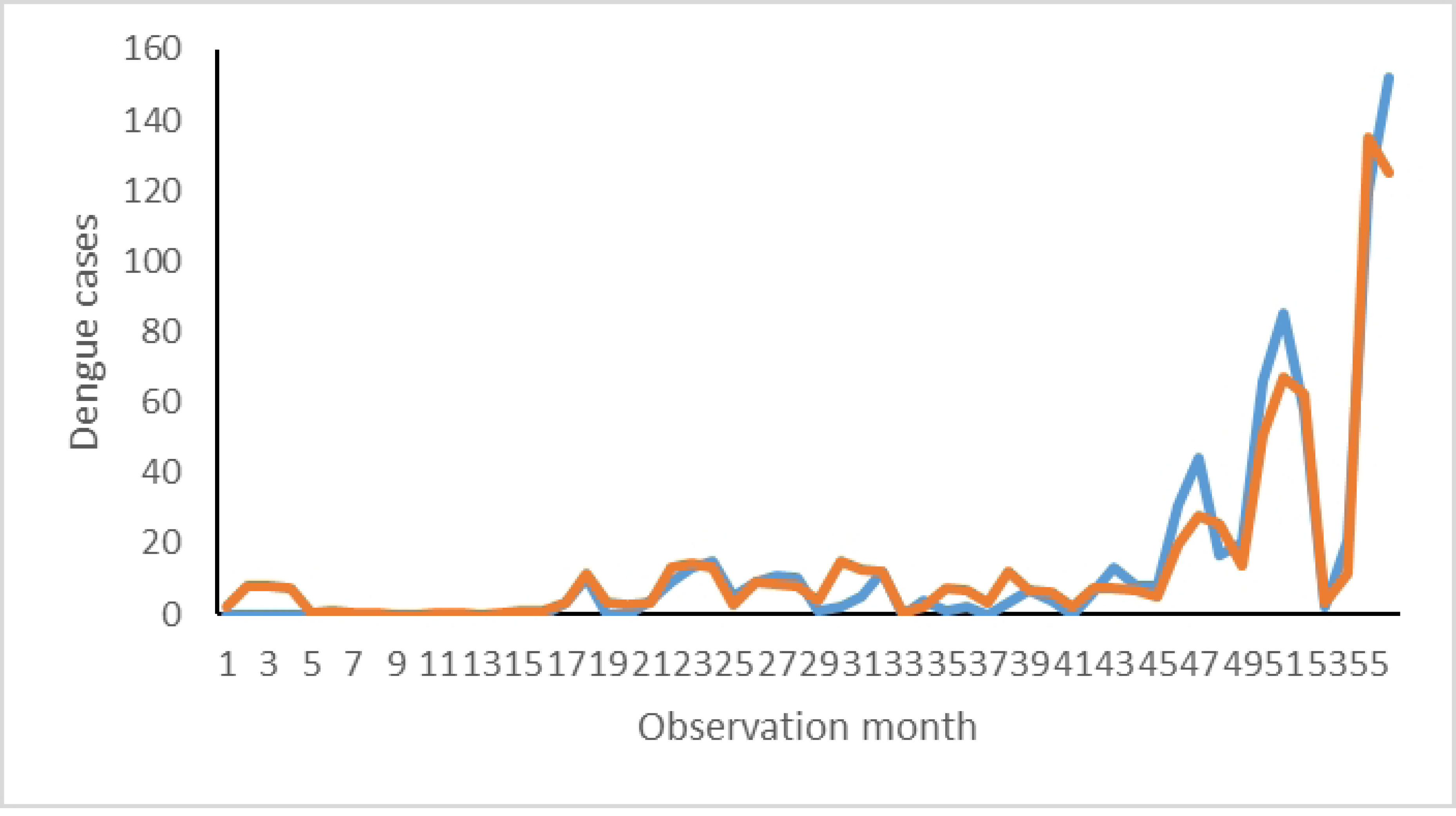
Fitted model of meteorological variables with the observed number of dengue cases during the same month in Luzon. Observed values shown by the blue line and fitted values by the orange line. The X-axis is the observation month for each of the two study sites starting from the beginning of the study until the end. Thus months 1-4 will be the first observation months of Cupang, Putatan, Sampaloc and Tambunting respectively and so forth.

### Association of mosquito indices with dengue cases

Whilst Container, House and Breteau indices and cumulative adult mosquitoes (over 2 months) were all associated with dengue cases in the same month in the univariable analyses, only the latter was positively associated with dengue cases in the final multivariable analysis (t=3.32, P=0.001), explaining 4.79% of the variation in the number of dengue cases (Figure S19). There was an increased Relative Risk of 1.52 (95%CI 1.18-1.95) for every increase in one mosquito in the standardized mosquito number, equivalent to a Relative Risk of 1.02 (95% CI 1.01-1.03) for every increase in one mosquito (unstandardized). This positive association was lost when combined with the meteorological variables. There were no associations of mosquito indices with dengue cases the following one or two months.

## Discussion

Meteorological conditions can influence the spread of dengue through their impact on the vector’s life cycle and ability to transmit the virus. This study aimed to expand the knowledge on the associations and their time lags between different meteorological variables and mosquito indices on the one hand and dengue incidence on the other. This study also aimed to assess the added value of incorporating the entomological indices in explaining variation in dengue incidence.

There were several non-linear associations between meteorological variables and the immature entomological indices, namely HI, CI, BI, PI and PPI. The most pertinent variables were those associated with rain (whether cumulative or mean daily rain over the month) and the DTR. These variables explained 49-63% of the variation in the immature indices. The rain variables showed a distinct plateau, with increasing indices up a certain level of precipitation but not beyond. Heavy rain has been suggested to result in a decrease in immature mosquito indices by flushing out the immature stages, thus decreasing its population and ability to transmit the disease [32, 34]. This study found no evidence of a decrease as might be expected if excess rain lead to flushing of the oviposition sites. The plateauing out of the indices with increasing rain might reflect a saturation in the number of available water-filled oviposition sites. Although the number of potential containers increases with population density, a modelling study on the relationship between human and mosquito densities has previously suggested that water container number likely does not keep increasing after a given number of humans is reached [35]. The non-linear relationship of immature indices with DTR is a well-recognised phenomenon whereby larval developmental time and time to pupation is impacted by higher DTR [36, 37]. Furthermore, Carrington et al. found that both small and large DTRs can affect the population dynamics [38].

In contrast to the immature indices, the adult mosquito numbers were most strongly influenced by minimum temperatures and Cumulative rain, with moderate effects of the other meteorological variables. No plateau in adult mosquito numbers was observed with increasing Cumulative rain, but which may reflect the weak correlation between immature and adult numbers. The association with minimum temperature most likely reflects the more general effects of temperature on adult longevity with peak survival rates occurring round 27°C and increased mortality rates occurring above 32°C [39-41]. The temperature variables are all highly correlated and thus the minimum temperature *per se* may not be the most biologically important.

The absence of any improved model fitting when using entomological indices lagged by one to four weeks after the meteorological variables suggests that the current month’s weather is that impacting the current month’s mosquito numbers. It should be noted, however, that although there was not an improved model fit, the meteorological variables did explain a substantial amount of the observed variation, ranging up to 64% for a four week lag with adult mosquitoes (as compared to 72% for the same month’s weather). This relatively strong association might be of value for an advanced warning index. In addition, the absence of improved lag effects may also reflect the relatively low variation in the meteorological variables over such a short time period and a longer time frame might be more informative, especially with respect to the adult population density following the dry season.

Similarly to previous studies, most meteorological variables had a non-linear association with the incidence of dengue [32, 42, 43]. Temperature fluctuations have an impact on the mosquito’s life span, development, reproduction rates, and feeding frequency, as well as the speed of virus replication and extrinsic incubation rate [19, 44]. Low temperatures have been associated with a decreased vector capacity, and here we clearly observed an accelerating risk of dengue beyond a minimum temperature of ∼22°C. High temperatures have also been found to be associated with decreased risk of dengue, generating a non-linear pattern [21, 32, 42, 45]. This is also observed in the present study, with a trend for decreased dengue incidence at temperatures beyond 33°C. A suggestive decrease in dengue risk was also observed beyond 34°C in Guangzhou [46]. Previous estimates of threshold temperature values for increased risk of dengue incidence, all from Guangzhou, China have identified values of ca. 18-23°C and 22-32°C for minimum and maximum temperature thresholds [46-48]. The values we found fall within the same range.

Lower diurnal temperature ranges have been associated with a higher spread of the dengue [20]. Moreover, less temperature variation during the day can also coincide with an optimal temperature mean for dengue transmission. A modelling approach found that decreased temperature variation around the predicted optimal temperature (∼29°C) increases the chances of viral transmission [21]. This negative association between temperature variation and dengue incidence was observed here and found only to lead to reduced risk at DTR above 6.5°C. Below this DTR, mean temperatures ranged from 27-29°C, suggesting an optimal combination of temperatures for transmission.

Rainfall can increase dengue transmission, generating more abundant oviposition sites and maintaining a sufficient relative humidity for adult mosquito survival [19]. A positive association with cumulative rainfall was observed here, with incidence increasing with as little as 100mm rainfall in a month. The similarity in the shape of the relationship of rain with adult mosquitoes and rain with dengue cases would suggest that this latter is occurring through the increase in adult mosquito numbers. This is supported by the observation of a positive association of cumulated adult mosquito numbers and dengue incidence.

When analysing the study sites on the island of Palawan separately from those on Luzon led to vastly improved model fit, especially for dengue cases occurring two months later. The same occurred when analysing the Luzon sites separately from the Palawan sites, but no added value was observed when analysing each city within Luzon separately. It is tempting to hypothesise that the weather variables are differentially affecting the two *Aedes* species that have differing relative abundances on the two islands with knock-on effects for explaining variation in dengue incidence. That the significantly associated meteorological variables remained over one to two months lag time is consistent with previous findings and potentially useful for any advanced warning strategy [32, 42]. Such long delayed effects are biologically plausible, reflecting the delay between amplification of the mosquito population, initial spread and subsequent expansion of the viral population. DENV has a lifecycle taking a minimum of 15 days, including a ∼10 day extrinsic incubation period within the mosquito following an infective bloodmeal and then a 4-10 day intrinsic incubation period following an infectious bite on a human. Two months lag would thus correspond to a maximum of three generations of viral transmission even with an adequate mosquito population density. Furthermore, given that the majority of DENV infections are inapparent, the consequences of expanding viral circulation identified through clinical cases would take longer [49].

There are several limitations to this study. Firstly, we only had 14 months dengue case data upon which to base our analyses, but which was carried out in six sites on two environmentally differing islands. Secondly, even though we had meteorological data from each of the cities, we had no more specific information at the scale of each site and thus could not include a spatial element in the analyses. Intra-annual spatial variation in incidence was observed, but may more likely reflect the known clustered nature of dengue outbreaks and the impact of herd immunity [50, 51]. Whilst meteorological variables likely vary at very local scales, disentangling the relative roles of immunity, clustering and meteorology will require fine-scale measures of serology, meteorology and case geolocalisation.

In conclusion, there have been increasing efforts to establish early warning and response systems using meteorological information, but which although promising have often proven country-specific [31, 32, 52]. The clear associations of meteorological variables and the mosquito indices is encouraging and yet it seems that the link between such indices and dengue incidence remains non-significant. Only adult mosquitoes showed any positive association, but far weaker than when using meteorological variables. In short, given the considerable effort required to obtain such entomological indices and their lack of any added value for predicting dengue outbreaks, further emphasis on meteorological variables at a more local scale might be a more promising route of investigation.

## Data Availability

Data are freely available on Figshare private link: https://figshare.com/s/0c5efc295eac6a3f09b5 and doi: 10.6084/m9.figshare.23978448

https://dx.doi.org/10.6084/m9.figshare.23978448

## Supplementary Information

**Figure S1** Mean Daily Rain (mm) by month in each of the three cities. Shown are means and standard errors of the mean.

**Figure S2** Mean Daily Relative Humidity (%) by month in each of the three cities. Shown are means and standard errors of the mean.

**Figure S3** Mean Daily Maximum Temperature (°C) by month in each of the three cities. Shown are means and standard errors of the mean.

**Figure S4** Mean Daily Minimum Temperature (°C) by month in each of the three cities. Shown are means and standard errors of the mean.

**Figure S5** Mean Daily Mean Temperature (°C) by month in each of the three cities. Shown are means and standard errors of the mean.

**Figure S6** Mean Daily Diurnal Temperature Range (°C) by month in each of the three cities. Shown are means and standard errors of the mean.

**Figure S7** House Indices per month in the six study sites. Shown are % and 95% Confidence Intervals.

**Figure S8** Fitted logistic regression model of House Index (here given as a proportion) against Cumulative Rain during the month. Red line shows the model output for the association of cumulative rain with HI and the actual data are crosses.

**Figure S9** Container Indices per month in the six study sites. Shown are % and 95% Confidence Intervals.

**Figure S10** Breteau Indices per month in the six study sites.

**Figure S11** Pupal Indices per month in the six study sites.

**Figure S12** Pupa per Person Indices per month in the six study sites.

**Figure S13** Number of adult Aedes spp. per month in the six study sites.

**Figure S14** The fitted loglinear regression of the number of adult Aedes spp. mosquitoes against the monthly Cumulative rain. The red line shows the fitted model in the GAM.

**Figure S15** The fitted loglinear regression of the number of adult Aedes spp mosquitoes against the monthly mean minimum temperature. The red line shows the fitted model in the GAM.

**Figure S16** The fitted loglinear regression of the lagged 2 week Pupal Index against the monthly mean Diurnal Temperature range. The red line shows the fitted model in the GAM.

**Figure S17** The fitted loglinear regression of the lagged 2 week Pupal Index against the monthly cumulative rain. The red line shows the fitted model in the GAM.

**Figure S18** The fitted loglinear regression of the mean Diurnal temperature range against the number of dengue cases the same month. The red line shows the fitted model in the GAM.

**Figure S19** The fitted loglinear regression of the standardized cumulative adult mosquitoes against the number of dengue cases the same month. The red line shows the fitted model in the GAM.

**Table S1** Association of meteorological variables with lagged mosquito indices. For each index we do 4 (lag weeks) ×7 (meteorological variables) univariable analyses plus 1 multi per lag week so 29. Bonferroni correction P value = 0.0017. P values in italics are those above this P value threshold.

**Data reporting Data are freely available on Figshare private link:** https://figshare.com/s/0c5efc295eac6a3f09b5 **and** doi: 10.6084/m9.figshare.23978448

